# The Moderating Role of Family Poverty-to-Income Ratio in the Depression-Mortality Association: A 2005-2018 NHANES Cohort Study

**DOI:** 10.1101/2025.06.05.25329093

**Authors:** Fang Yao, Jiayao Zhang, Hui Wang

## Abstract

**Background:** To investigate the mediating role of family poverty income ratio (PIR) in the association between depression and mortality risk, including all-cause and cardiovascular disease mortality, using data from the National Health and Nutrition Examination Survey (NHANES).

**Methods:** The study included a cohort of participants from seven NHANES cycles (2005–2018) with available depressive assessments and family PIR data. The primary endpoint was all-cause mortality, and the secondary endpoint was cardiovascular mortality, ascertained from National Center for Health Statistics (NCHS) mortality follow-up data. To evaluate the relationship between depression and mortality, we employed restricted cubic splines (RCS), Cox proportional hazards regression models. After adjusting for covariates such as age, sex, education, and marital status, mediation analysis was conducted to assess the mediating effects of the family PIR on these associations.

**Results:** The prevalence of depressive symptoms among the 33,132 participants was 8.64%, with a mean age of 47.62 ± 18.69 years. After adjusting for multiple confounders, higher Patient Health Questionnaire-9(PHQ-9) scores were significantly associated with an increased risk of all-cause mortality (HR = 1.050, 95% CI: 1.042–1.058, *P* < 0.0001) and cardiovascular mortality (HR = 1.035, 95% CI: 1.019–1.050, *P* < 0.00001). Mediation analysis revealed that 14.51% and 5.82% of the effects of depression on all-cause and cardiovascular mortality, respectively, were mediated through family PIR.

**Conclusion:** The family PIR plays a partial mediating role in the relationship between depressive and mortality risk, highlighting the importance of socioeconomic factors in understanding and reducing depression-related mortality. Interventions targeting both mental health and socioeconomic support are essential to improve overall survival among individuals with depression.

## 1. Introduction

Depression is a significant global mental health concern, affecting approximately 280 million individuals worldwide, making it a leading cause of disability. The lifetime prevalence of major depressive disorder is estimated to range between 10% and 15%, while the 12-month prevalence is reported at approximately 4.4%[1]. Notably, during significant events such as the COVID-19 pandemic, the prevalence of depression experienced a marked increase, with some estimates reaching as high as 24%[2]. The multifactorial nature of depression means that its impact on health is profound, contributing not only to individual suffering but also to a substantial increase in all-cause mortality risk. Research indicates that the relationship between depression and mortality can be mediated through various behavioral, biological, and social pathways [3–5]. For instance, individuals suffering from depression may engage in unhealthy behaviors such as reduced physical activity, poor dietary choices, and substance abuse, which can exacerbate physical health problems and lead to premature death[6–8] .

Despite the clear association between depression and increased mortality, the precise nature of this relationship warrants further exploration. Multiple studies have highlighted that the effects of depression on mortality rates can vary based on demographic factors, including age, gender, and the presence of chronic illnesses [9–11]. For example, men typically have a heightened risk of mortality linked to depression compared to women, possibly due to differences in help-seeking behaviors and social support systems [12, 13]. Additionally, some meta-analyses have revealed that while depression consistently shows a correlation with higher mortality rates, this association may diminish when controlling for various confounding factors such as socioeconomic status and baseline health conditions [14, 15].

Socioeconomic status (SES) remains a critical determinant of mental health outcomes, particularly in relation to depression and mortality risk. The family poverty-to-income ratio (PIR) serves as a standardized measure of SES, illustrating the precarious position of families living below a defined poverty threshold. Lower PIR values correlate strongly with increased socioeconomic disadvantages, manifesting in heightened risks for both depression and premature mortality[16]. A study by Lorant et al. emphasizes this correlation, indicating that individuals from lower SES backgrounds experience a marked increase in depressive symptoms, further exacerbated by age and gender dynamics within the population under study[17]. Additionally, research by Lund et al. details how poverty creates a “vicious cycle”intensifying the prevalence of common mental disorders and limiting individuals’ capacity to access necessary mental health services[18].

Moreover, recent evidence suggests that the mediating role of socioeconomic disadvantage in the health impacts of depression warrants further exploration. For example, Flèche and Layard elaborate on how mental pain can be a significant barrier to adapting successfully to socioeconomic challenges, arguing for a more nuanced understanding that encompasses both mental health and socioeconomic conditions as intertwined factors impacting overall health outcomes[19]. The acknowledgment that social capital can play a role in mediating these effects suggests that interventions aimed at improving community ties and welfare may offer pathways to mitigate the adverse health effects associated with poverty [20, 21]. In summary, the complex interplay between socioeconomic status, depression, and health outcomes necessitates a comprehensive approach that considers both subjective experiences of poverty and objective measures like The family PIR. Addressing these concerns holistically may enhance mental health outcomes and reduce mortality risks among disadvantaged groups.

The aim of this study was to conduct a cohort analysis using data from the National Health and Nutrition Examination Survey (NHANES) collected between 2005 and 2018, in order to investigate the mediating role of the family PIR in the association between depression and mortality.

## 2. Methods

### 2.1 Participants

This study utilized data from seven cycles (2005-2006, 2007-2008, 2009-2010, 2011-2012, 2013-2014, 2015-2016, and 2017-2018) of the National Health and Nutrition Examination Survey (NHANES) database conducted by the National Center for Health Statistics (NCHS) under the Centers for Disease Control and Prevention (CDC). NHANES employs a stratified multistage probability sampling design to systematically collect health and nutrition data from the non-institutionalized civilian population of the United States. Written informed consent was obtained from all participants, and the study protocol was approved by the NCHS Ethics Review Committee. This was a nationally representative cohort study based on NHANES participants. The study excluded participants who were younger than 18 years of age (n=28,047), those with missing depression data (n=37,012), those with missing mortality data (n=66), and those with missing Family PIR data (n=3,061). The final sample included 33,132 individuals for analysis. The process of participant inclusion and exclusion is illustrated in Fig 1.

**Fig. 1.**
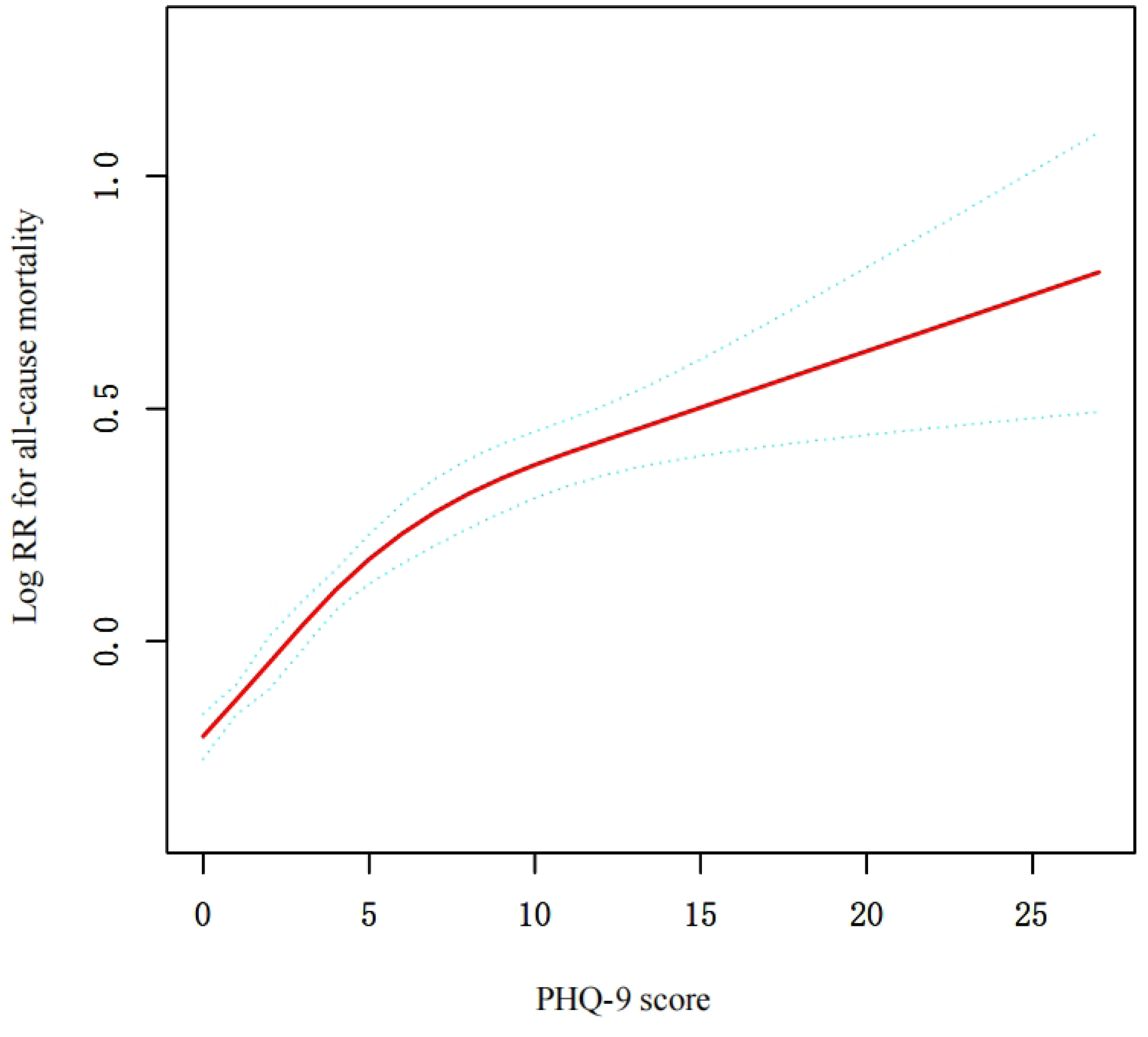
Participant selection flowchart for NHANES study (2005–2018)

### 2.2 Measurements

#### 2.2.1 Depression

The assessment of depressive symptoms was conducted using the Patient Health Questionnaire-9 (PHQ-9), a self-report questionnaire that is based on the nine diagnostic criteria for depression that are listed in the Diagnostic and Statistical Manual of Mental Disorders, Fourth Edition (DSM-IV)[22]. The PHQ-9 comprises nine items, with each question scored on a scale ranging from “0” (not at all) to “3” (nearly every day), resulting in a total score ranging from 0 to 27. The questionnaire demonstrates strong reliability and validity, with a Cronbach’s α of 0.89[23], thus confirming its psychometric properties across diverse populations. The establishment of a total score of 10 as the clinical diagnostic threshold is congruent with the DSM-IV criteria for depression. Scores of 10 or higher are considered indicative of clinically significant depressive symptoms[24].

#### 2.2.2 All cause mortality

The primary outcome assessed was all-cause mortality,with follow-up information collected via linkage to the National Death Index (NDI)[25], up to December 31, 2019. Cardiovascular disease (CVD) mortality was classified based on the Tenth Revision of the International Statistical Classification of Diseases and Related Health Problems (ICD-10) criteria, including (1)cerebrovascular diseases (I60-I69) and (2) diseases of the heart (I00-I09, I11, I13, I20-I51).

#### 2.2.3 Family Poverty-to-Income Ratio

Socioeconomic status was evaluated by the Poverty Income Ratio (PIR), which is determined by dividing a family’s income by the established poverty threshold[26].

#### 2.2.4 Covariates

We included a number of covariates that may have influenced the results: age, sex, race, educational level (incomplete 9th grade/9th-11th grade (including 12th grade without diploma)/high school graduate/partial college or associate’s degree/college graduate and above) and marital status (married/living with partner/never married/divorced/separated/widowed), body mass index (BMI) (<25 ,25-30, ≥30)[27,28]. These covariates were collected using a standardised questionnaire. Weight and height of each participant were determined by physical examination. Body mass index (BMI) was defined as weight (kg) divided by height squared (m). The number of missing values for the covariates among the 33,132 patients was 1934 (5.84%) for education level, 1448 (4.37%) for marital status, 341 (1.03%) for BMI and 30(0.09%)for asthma history.

### 2.3 Statistic analysis

All statistical analyses were performed using EmpowerStats software (www.empowerstats.com, X&Y Solutions, Inc. Boston, MA) and R software version 4.2.0 (http://www.r-project.org). All statistical tests were two-sided, and a *P* value < 0.05 was considered statistically significant.

First, the baseline characteristics of the study population were described.Continuous variables are reported as mean values with standard deviations (mean ± SD), whereas categorical variables are presented as frequencies and percentages [n (%)]. Participants were grouped according to the presence or absence of depression, and group differences were compared. The comparison of continuous variables was performed using the Mann–Whitney U test, while the comparison of categorical variables was conducted using the Chi-square test.

Next, restricted cubic spline and Cox proportional hazards regression models were used to evaluate the association between baseline PHQ-9 depression scores and the risk of all-cause mortality. If a significant nonlinear relationship was identified in the spline model, a piecewise Cox regression model was further applied, and the likelihood ratio test was used to compare the goodness-of-fit between the linear and piecewise models. A *P* value < 0.05 was considered indicative that the piecewise model fit the data better, suggesting a statistically significant threshold effect. Survival analysis was conducted using the Kaplan–Meier method to estimate survival probabilities for depressed and non-depressed groups, and the log-rank test was employed to compare survival curves between the two groups.

To evaluate the independent association between baseline PHQ-9 depression scores and the risks of all-cause and cardiovascular mortality, three Cox proportional hazards models were constructed: Model 1 was unadjusted; Model 2 was adjusted for sex, age, and race; and Model 3 was further adjusted for education level, marital status, body mass index (BMI), and history of asthma based on Model 2.Additionally, linear regression and Cox proportional hazards regression models were employed to evaluate the associations of Family PIR with depression and with mortality, respectively. For each association, unadjusted, partially adjusted, and fully adjusted models were constructed.

Finally, mediation analysis was conducted using the“ mediation ” package in R software (version 4.2.0) to assess the mediating effect of family PIR on the relationship between depression and mortality[29]. This analysis adjusted for confounding factors, including sex, age, race, education level, marital status, BMI, and history of asthma.

## 3. Results

### 3.1 Baseline characteristics

Table 1 presents the baseline characteristics of 33,132 participants regarding depressive symptoms and all-cause mortality. The prevalence of depressive symptoms in the study population was 8.64% (2,867/33,132), with a mean age of 47.624 ± 18.692 years, and females accounted for 50.82% of the participants. Individuals with depressive symptoms reported significantly lower family PIR compared to those without depression (1.706 ± 1.372 vs. 2.565 ± 1.630, *P* < 0.001), and had a notably higher average body mass index (30.836 ± 8.429 kg/m² vs. 28.937 ± 6.885 kg/m², *P* < 0.001). The proportion of females in the depressive group was significantly higher than that of males (63.52% vs. 36.48%, *P* < 0.001), and the educational level was lower, with only 10.56% holding a university degree or higher. Furthermore, the all-cause mortality rate in the depressive group was 12.243%, significantly exceeding that of the non-depressive group (9.767%, *P* < 0.001).

**Table 1.**
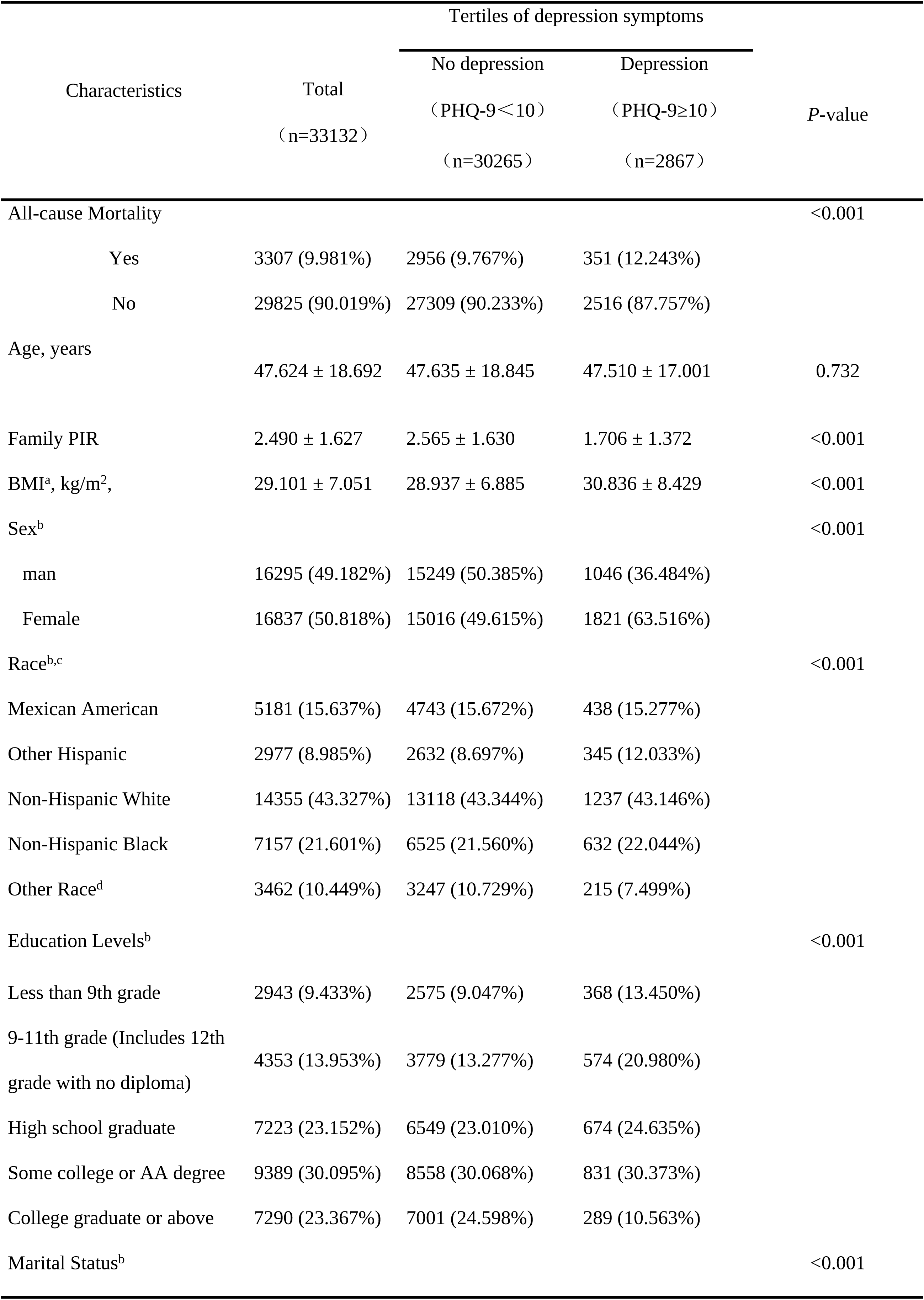
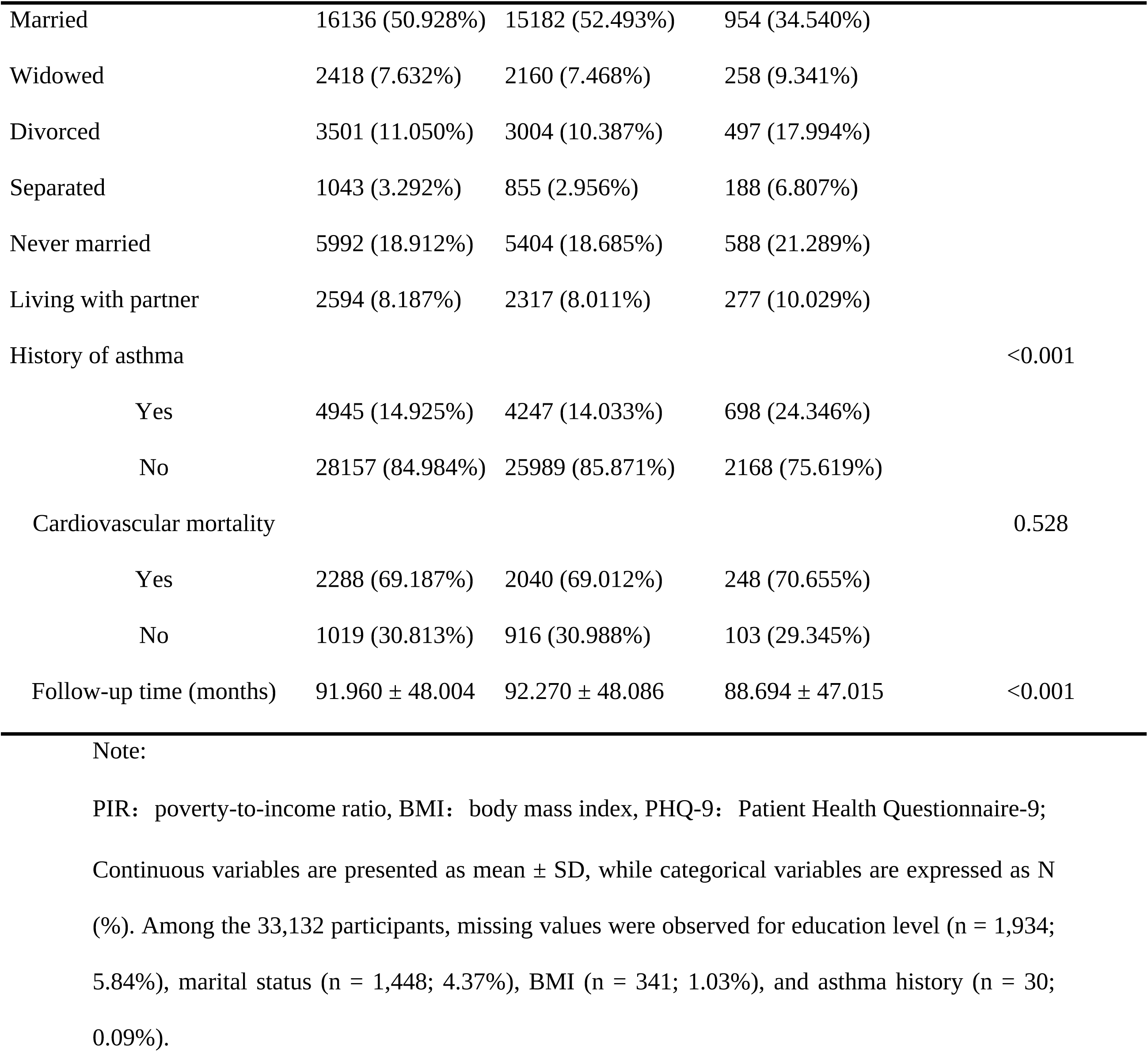
Baseline Characteristics of the Study Population Grouped by Depression Status

### 3.2 Association between depression and all-cause mortality

The association between PHQ-9 score and all-cause mortality was evaluated using a two-piecewise linear regression model, as depicted in Fig 2 and Table 2. The results demonstrated a significant positive association between PHQ-9 score and the logarithm of the relative risk (Log RR) for all-cause mortality, with the relationship exhibiting a non-linear trend. Model 1 indicated that every one-unit increase in PHQ-9 score was associated with a 5.0% increased risk of all-cause mortality (HR=1.050, 95% CI: 1.042–1.058, *P* <0.0001). Model 2 identified an inflection point at a PHQ-9 score of 7, showing a stronger association below this threshold (HR = 1.078, 95% CI: 1.061–1.095, *P* < 0.0001) compared to scores above 7 (HR = 1.021, 95% CI: 1.005–1.038, *P* = 0.0120). The log-likelihood ratio test indicated that the two-piecewise linear regression model provided a significantly better fit for the data (*P* < 0.001).

**Fig. 2.**
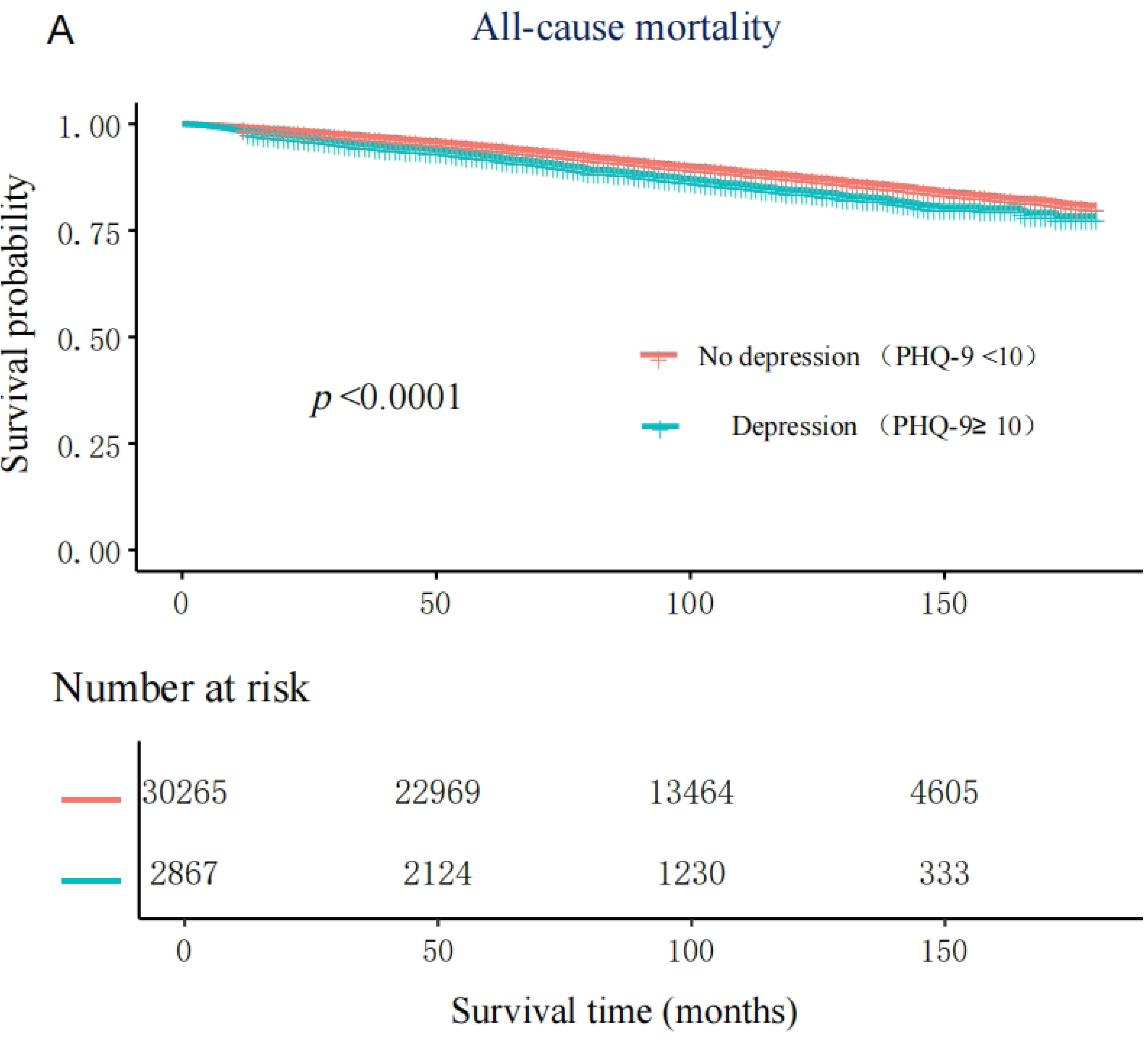
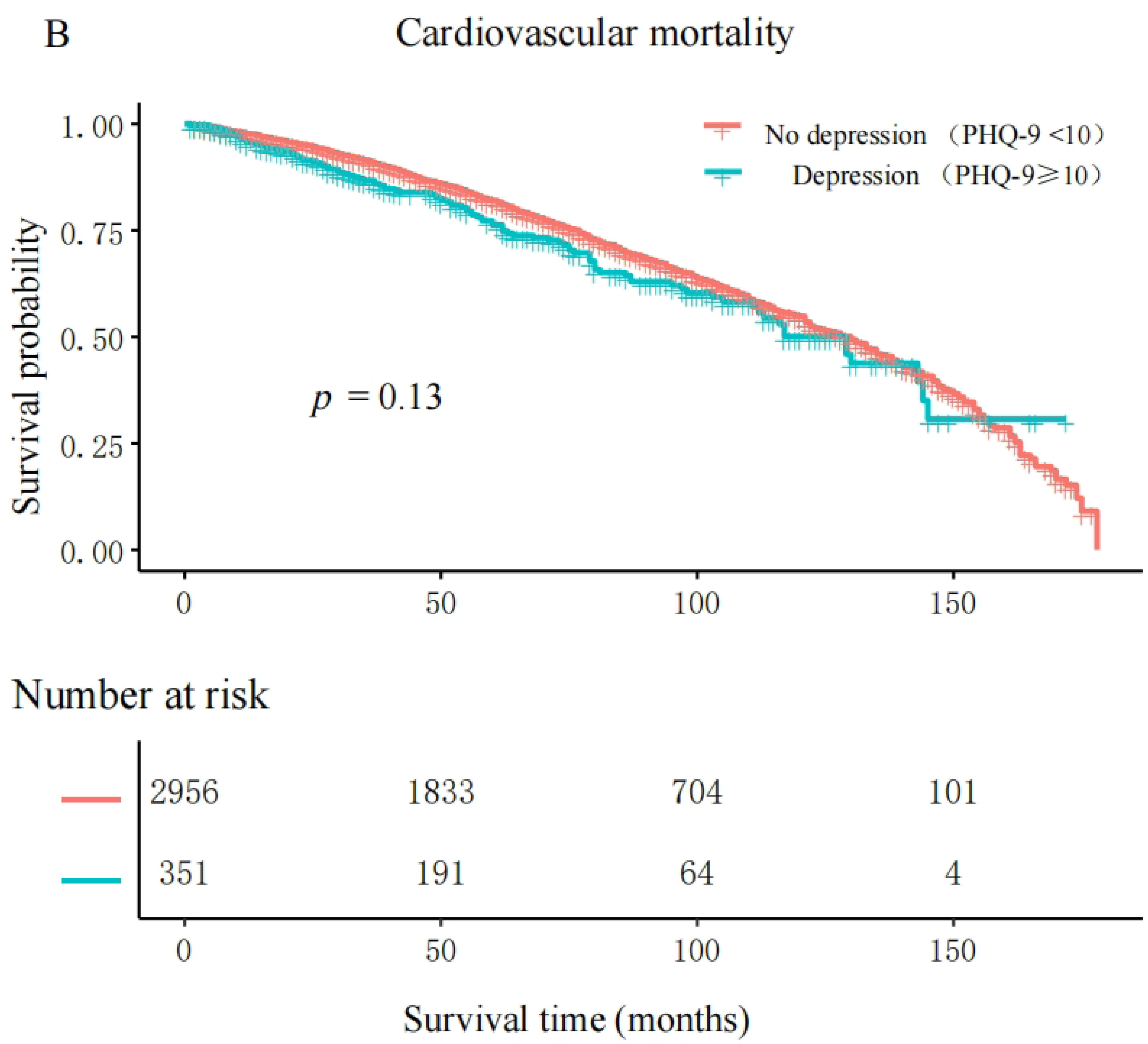
Dose-response relationship between PHQ-9 Score and All-Cause Mortality. Sex, age, race, education levels, marital status, BMI, history of asthma were adjusted.

**Table 2.**
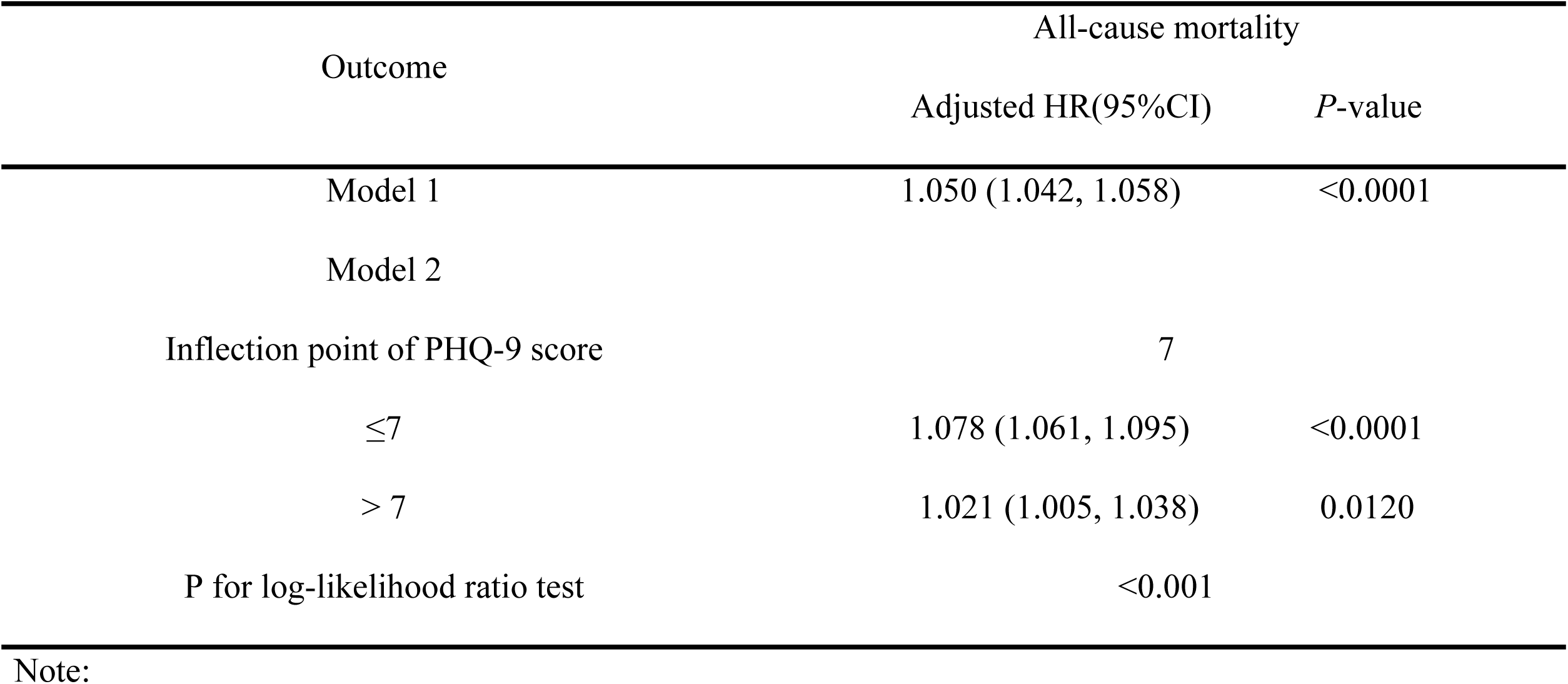
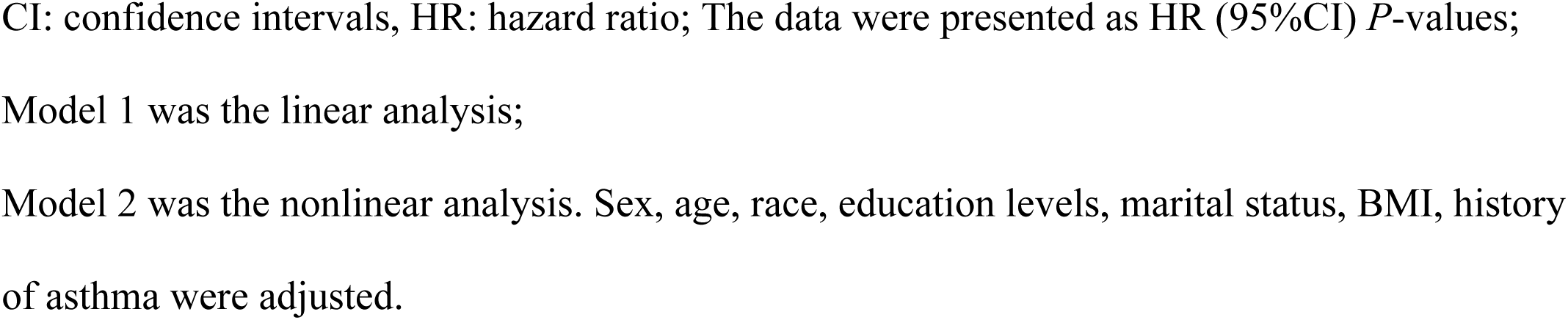
Threshold effect analysis of depression and all-cause mortality

| Outcome | All-cause mortality |  |
| --- | --- | --- |
|  | Adjusted HR(95%CI) | *P*-value |
| Model 1 | 1.050 (1.042, 1.058) | <0.0001 |
| Model 2 |  |  |
| Inflection point of PHQ-9 score | 7 |  |
| ≤7 | 1.078 (1.061, 1.095) | <0.0001 |
| > 7 | 1.021 (1.005, 1.038) | 0.0120 |
| P for log-likelihood ratio test | <0.001 |  |

Note:
CI: confidence intervals, HR: hazard ratio; The data were presented as HR (95%CI) *P*-values; Model 1 was the linear analysis; Model 2 was the nonlinear analysis. Sex, age, race, education levels, marital status, BMI, history of asthma were adjusted.

Kaplan–Meier survival curves were constructed for participants with depression (PHQ-9 ≥ 10) and those with no depression (PHQ-9 < 10), and group differences were evaluated with the log-rank test. The depression group exhibited a significantly lower survival probability for all-cause mortality (Fig. 3(A); log-rank *P* < 0.0001), whereas no significant difference was observed between the two groups for cardiovascular mortality (Fig. 3(B); log-rank *P* = 0.13).

**Fig. 3.**
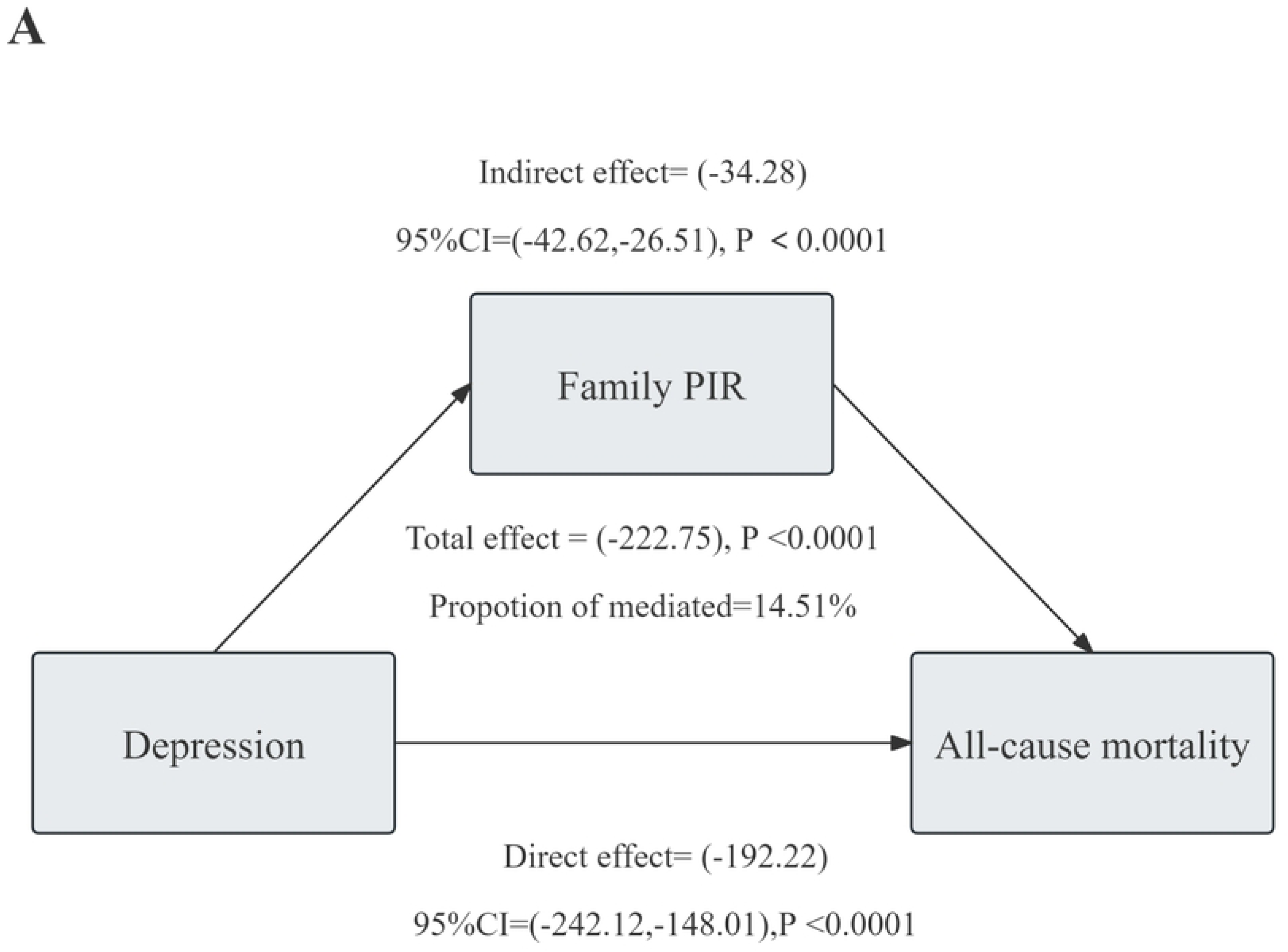
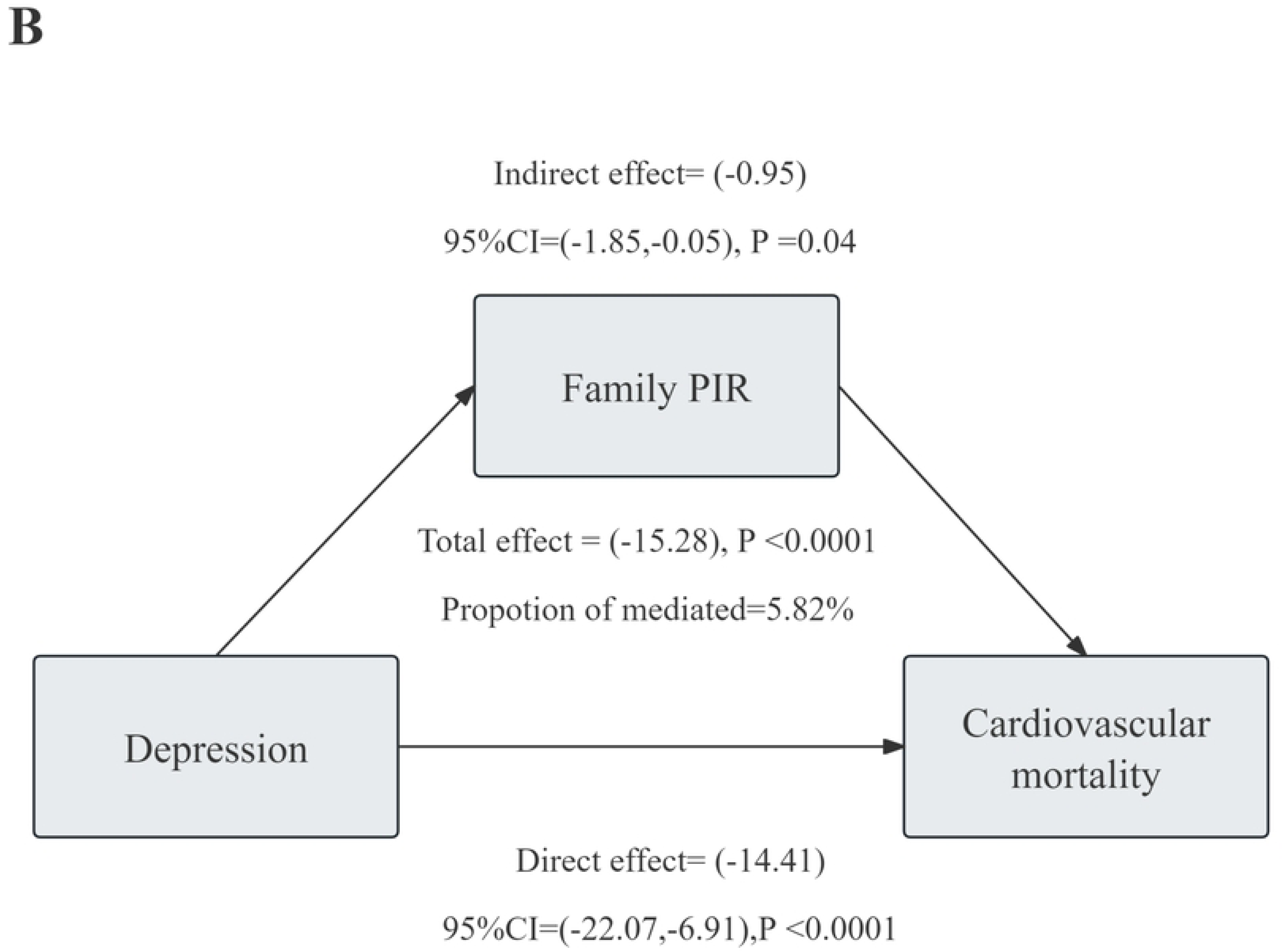
Kaplan-Meier survival analysis(A. All-cause mortality, B. Cardiovascular mortality)

Table 3 shows that depressive symptoms, as measured by PHQ-9 scores, are significantly positively associated with both all-cause and cardiovascular mortality. After adjustment in different models, each 1-point increase in PHQ-9 score was associated with a 5.0% higher risk of all-cause mortality in the fully adjusted Model 3 (HR = 1.050, 95% CI: 1.042–1.058, *P* < 0.00001), and a 3.5% higher risk of cardiovascular mortality (HR = 1.035, 95% CI: 1.019–1.050, *P* < 0.00001). These associations remained significant even after adjustment for multiple potential confounders.

**Table 3.**
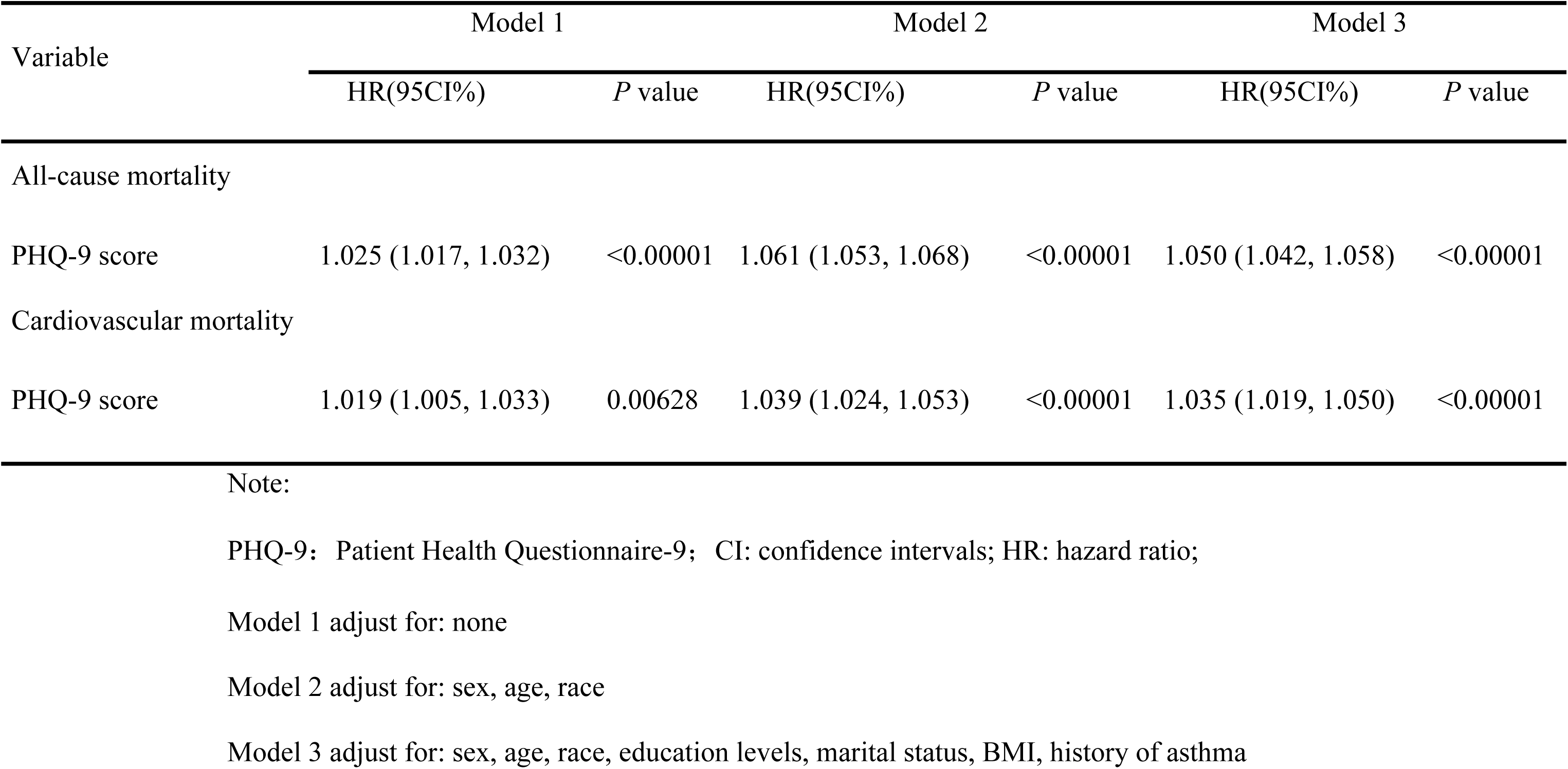
Crude and adjusted associations between depression and all-cause mortality and cardiovascular mortality in different models

### 3.3 Association of family PIR with depression and mortality

Table 4 revealed a significant association between depression, as measured by the PHQ-9 score, and the family PIR. A higher PHQ-9 score was consistently associated with a lower family PIR, and this relationship remained robust even after adjusting for potential confounding variables such as sex, age, race, education levels, marital status, BMI, and history of asthma. In the fully adjusted model, each unit increase in PHQ-9 score was associated with a significant decrease in family PIR (β= -0.051;95% CI: -0.054, -0.047; *P* < 0.000001). Furthermore, participants with depression (PHQ-9 scores ≥ 10 ) had a significantly lower family PIR compared to those with no depression (PHQ-9 scores < 10 ) (β= -0.583 ; 95% CI: -0.640, -0.527; *P* < 0.000001). These findings indicate that poverty, as measured by family PIR, has an independent and significant association with depression.

**Table 4.**
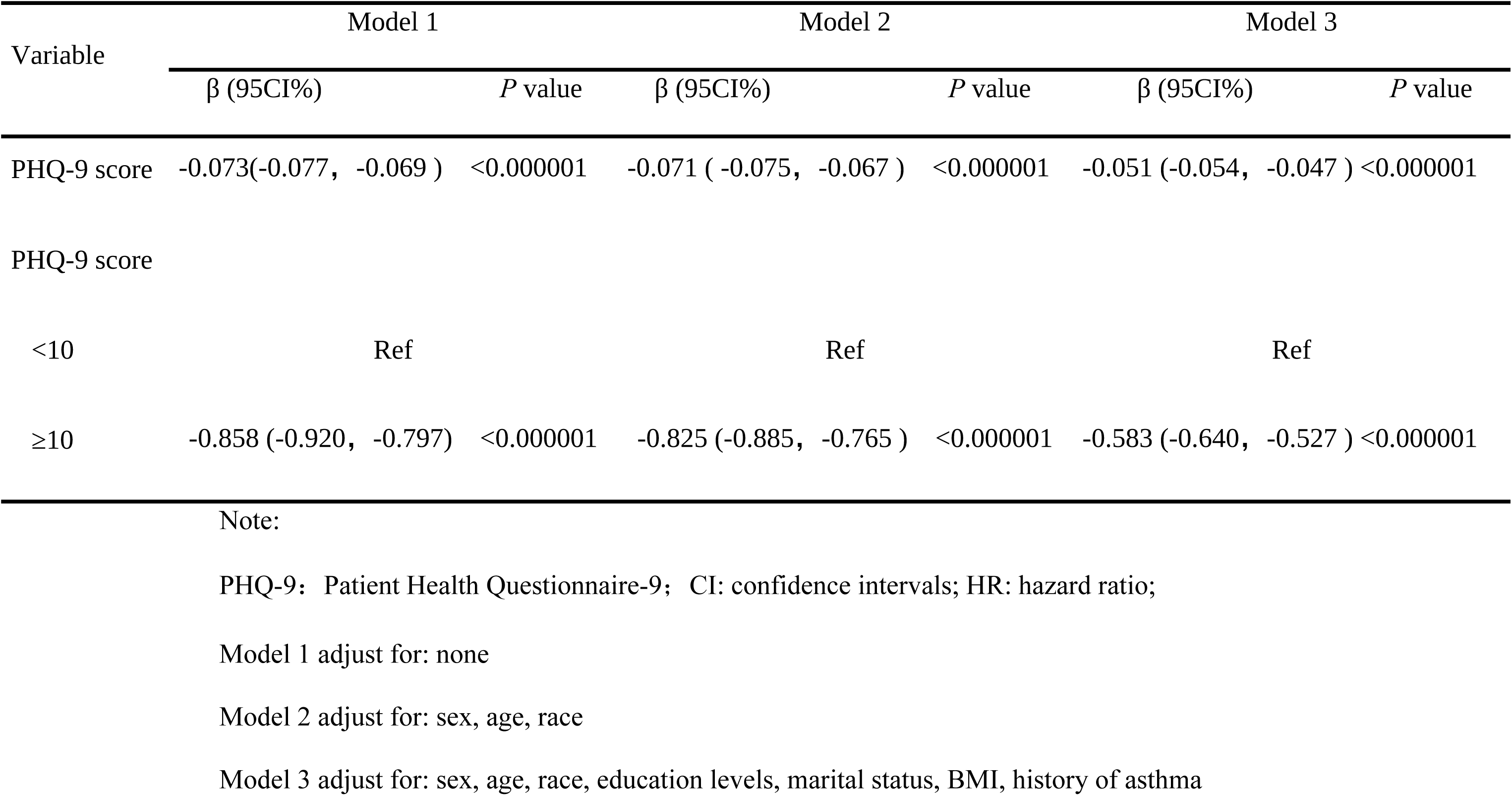
Crude and adjusted association between depression and family PIR in different models.

| Variable | Model 1 |  | Model 2 |  | Model 3 |  |
| --- | --- | --- | --- | --- | --- | --- |
| | $\beta$ (95CI%) | *P* value | $\beta$ (95CI%) | *P* value | $\beta$ (95CI%) | *P* value |
| PHQ-9 score | -0.073(-0.077, -0.069 ) | <0.000001 | -0.071 ( -0.075, -0.067 ) | <0.000001 | -0.051 (-0.054, -0.047 ) | <0.000001 |
| PHQ-9 score |  |  |  |  |  |  |
| <10 | Ref |  | Ref |  | Ref |  |
| $\geq 10$ | -0.858 (-0.920, -0.797) | <0.000001 | -0.825 (-0.885, -0.765 ) | <0.000001 | -0.583 (-0.640, -0.527 ) | <0.000001 |

Note: PHQ-9: Patient Health Questionnaire-9; CI: confidence intervals; HR: hazard ratio; Model 1 adjust for: none Model 2 adjust for: sex, age, race Model 3 adjust for: sex, age, race, education levels, marital status, BMI, history of asthma

Table 5 showed the association between family PIR and mortality (all-cause mortality and cardiovascular mortality). In both Model 1 (unadjusted) and Model 2 (partially adjusted for confounders), family PIR was negatively associated with all-cause and cardiovascular mortality. Even after adjusting for all confounders in Model 3, family PIR remained significantly negatively associated with all-cause mortality (HR = 0.831; 95% CI: 0.807–0.855; *P* < 0.00001) and cardiovascular mortality (HR = 0.930; 95% CI: 0.882–0.981; *P* = 0.00720).

**Table 5.**
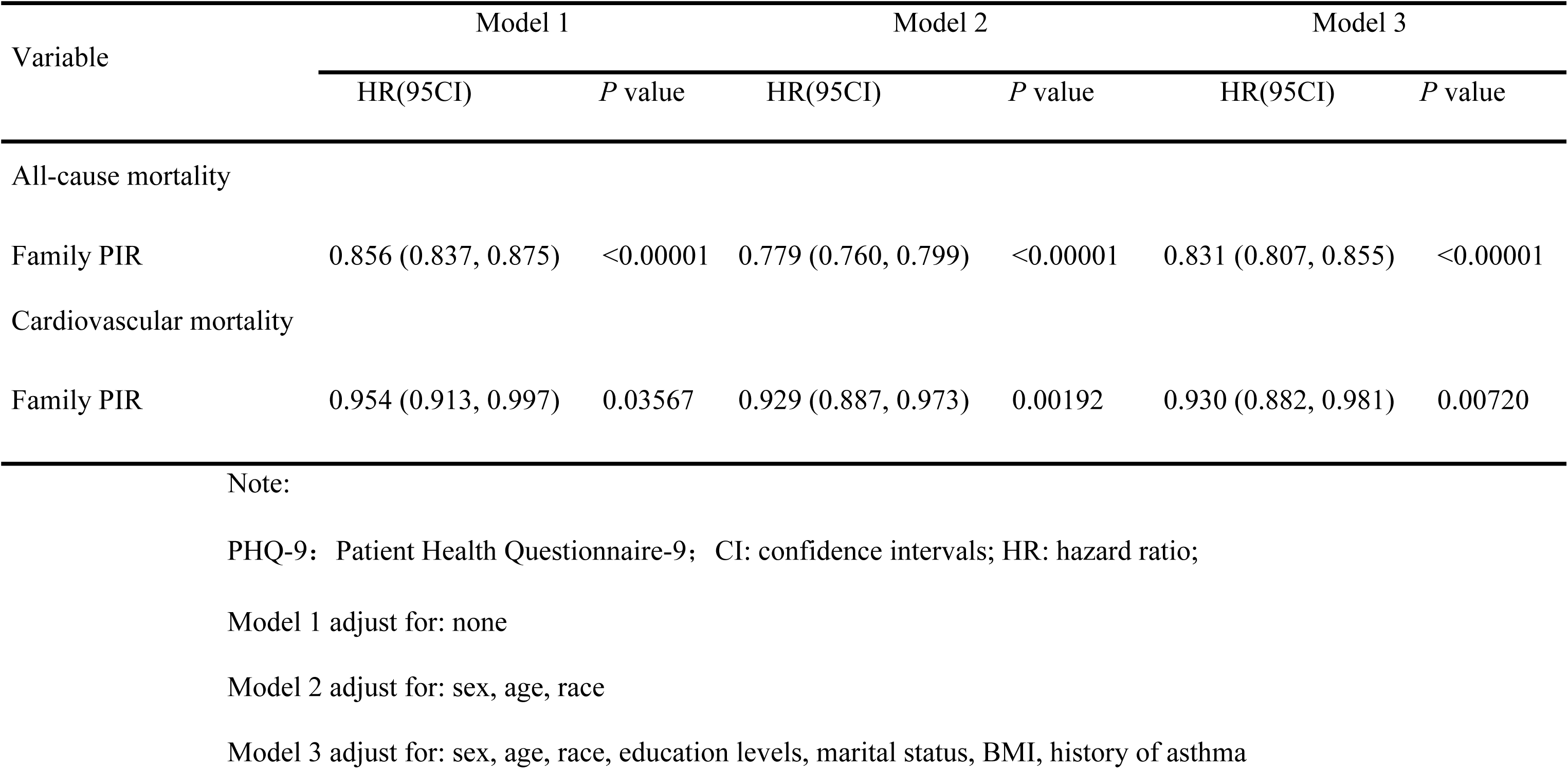
Crude and adjusted associations between family PIR and all-cause mortality and cardiovascular mortality in different models.

| Variable | Model 1 |  | Model 2 |  | Model 3 |  |
| --- | --- | --- | --- | --- | --- | --- |
|  | HR(95CI) | *P* value | HR(95CI) | *P* value | HR(95CI) | *P* value |
| All-cause mortality |  |  |  |  |  |  |
| Family PIR | 0.856 (0.837, 0.875) | <0.00001 | 0.779 (0.760, 0.799) | <0.00001 | 0.831 (0.807, 0.855) | <0.00001 |
| Cardiovascular mortality |  |  |  |  |  |  |
| Family PIR | 0.954 (0.913, 0.997) | 0.03567 | 0.929 (0.887, 0.973) | 0.00192 | 0.930 (0.882, 0.981) | 0.00720 |

Note:
PHQ-9: Patient Health Questionnaire-9; CI: confidence intervals; HR: hazard ratio;
Model 1 adjust for: none
Model 2 adjust for: sex, age, race
Model 3 adjust for: sex, age, race, education levels, marital status, BMI, history of asthma

### 3.4 Mediating role of family PIR

Fig.4 presents the mediation analysis examining the association between depression and mortality, with family PIR as a mediator. All analyses were adjusted for potential confounders, including sex, age, ethnicity, education level, marital status, body mass index, and history of asthma. As shown in Fig.4 (A), depression had a significant total effect on all-cause mortality (effect = -222.75, 95% CI: -242.12 , -148.01, *P* < 0.0001), with the indirect effect through family PIR accounting for 14.51% of the total effect (indirect effect = -34.28, 95% Cl: -42.62 to -26.51, *P* < 0.0001). For cardiovascular mortality (Fig.4 (B)), the total effect was also significant (effect = -15.28, 95% CI: -22.07 to -6.91, *P* < 0.0001), and the indirect effect via family PIR accounted for 5.82% (indirect effect = -0.95, 95% CI: -1.85 , -0.05, *P* = 0.04). In both models, the direct effects remained significant, suggesting that family PIR partially mediated the association between depression and mortality.

**Fig. 4.**
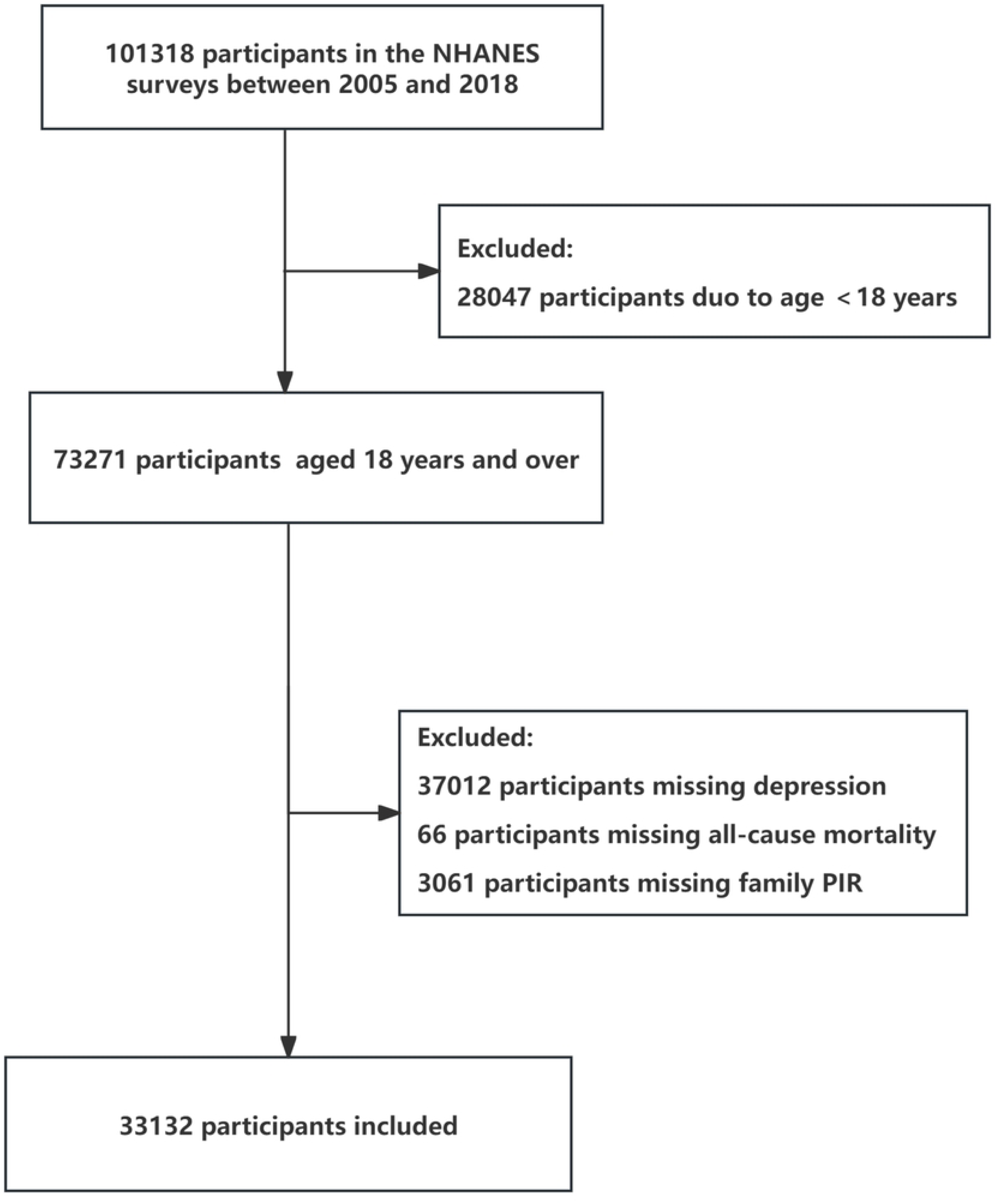
Mediation analysis of the association between depression and mortality through family PIR(A.All-cause Mortality; B. cardiovascular mortality). Sex, age, race, education levels, marital status, BMI, history of asthma were adjusted.

## 4. Discussion

The study analysed data from 33,132 participants in the U.S. National Health and Nutrition Examination Survey (NHANES) and NDI mortality data in order to investigate the relationships between famiy PIR, depression and mortality risk. The results demonstrated that higher PHQ-9 scores were significantly associated with an increased risk of all-cause mortality and cardiovascular mortality. Family PIR showed a negative correlation with depression and was also negatively associated with the risks of all-cause and cardiovascular mortality. Furthermore, depression mediated 14.51% of the association between family PIR and all-cause mortality, and 5.82% of the association between family PIR and cardiovascular mortality.

The findings of this study were consistent with previous literature [30,31]in several respects. Firstly, these studies all utilized the large-scale NHANES population database to systematically analyze the impact of depression on all-cause and cardiovascular mortality, and consistently confirmed that depression significantly increased the risk of these adverse outcomes. Specifically, a study involving 24,727 adult participants from the NHANES database demonstrated that social determinants of health (SDoH) played a moderating or mediating role in the relationship between depression and mortality risk ,and SDoH included factors such as family PIR (poverty income ratio)、education access and quality[30]. Similarly, another study with 28,439 respondents highlighted a strong association between socioeconomic status—including poverty and educational attainment—and both depression and mortality, underscoring the importance of considering socioeconomic variables when evaluating these associations [31]. Collectively, these findings emphasized that depression, as a significant public health issue, was closely related to socioeconomic background in both its occurrence and outcomes. Therefore, interventions targeting both psychological and socioeconomic determinants might be critical for reducing disparities in mortality among individuals with depression.

However, The present study focused on the family PIR as a singular economic indicator and quantified its mediating effect in the association between depression and mortality using mediation analysis, finding a mediation proportion of 14.5%. By contrast, Zun Wang et al. applied a composite Social Determinants of Health (SDoH) score, which yielded a stronger mediation effect (32.4%), indicating that methodological differences—specifically the choice and complexity of socioeconomic indicators—may have accounted for variations in mediation strength. In my study, family PIR, as a single economic indicator, was straightforward and easy to obtain. By focusing on economic factors, it helped reduce confounding influences. PIR directly reflected the economic status of households, making it practical for government and relevant departments to use in policy-making. This was beneficial for targeting and intervening in economically disadvantaged populations.

Additionally, different diagnostic tools were used. Laura A. Pratt’s study [31] utilized the CIDI-SF diagnostic tool, assessing anxiety and depression jointly, whereas the present study employed the PHQ-9, which specifically evaluated depressive symptoms. This difference in assessment instruments could have contributed to discrepancies in the detection rate and risk estimation of depression. Finally, A systematic review and meta-analysis reported that the association between depression and excess mortality among older adults could not be fully explained by economic factors, especially in low- and middle-income countries[32]. This contrasted with the findings of the current study, which were based on U.S. adults, likely due to differences in country-level economic status, health resources, and population structures. Therefore, variations in study populations, selection of socioeconomic measures, and depression assessment tools were key factors explaining the differences in results between the current study and previous literature, underscoring the need for context-specific and methodologically tailored research in this field.

This study found that depression was significantly associated with a higher risk of all-cause mortality and cardiovascular mortality, and revealed that family PIR partially mediated the relationship between depression and mortality. an integrated approach targeting mental health and socioeconomic factors can help to reduce mortality among people with depression.

The family PIR played a partial mediating role between depression and mortality, with its mechanisms manifesting in multiple dimensions. First, Economic pressure significantly increased the risk of depression by triggering chronic stress responses and weakened individuals’ ability to manage their mental health, which could have led to an increase in mortality[33–35].Research indicates that prolonged economic stressors, such as unemployment or reduced income, are linked to increased rates of anxiety and depression within populations, especially those already susceptible to mental health issues due to their socio-economic background[36–38]. A lower poverty-income ratio is typically accompanied by chronic economic stress, which manifests psychologically as sustained stress responses, elevates the risk of depression, and impairs disease management abilities—potentially leading to increased mortality [39, 40]. Second, economic stress exposes individuals to various life challenges, and numerous studies have shown a significant association between life stress and depression [41, 42].Furthermore, poverty limits access to social support and family resources, exacerbating feelings of loneliness and psychological burden[43]. Social support plays a critical role in alleviating depressive symptoms, particularly during times of financial hardship[44–46]. In the absence of such support, rates of loneliness and depression increase significantly [42, 47]. Research has demonstrated a direct link between lower levels of social support and higher incidence of depression; support from family and friends can effectively mitigate depressive symptoms[41]. Third, economic hardship often impedes individuals from maintaining healthy lifestyles and accessing essential healthcare resources, further increasing physical health risks[46, 48]. Financial constraints make it challenging to obtain nutritious foods and necessary medical services, contributing to a higher risk of various health problems, such as obesity and diabetes, which are closely related to family mental health[49, 50] .

It is noteworthy that exposure to poverty early in life can have long-term adverse effects on both psychological and physical health, ultimately increasing the risk of depression and mortality in adulthood[51].Individuals living in low socioeconomic status for extended periods experience persistent economic instability and chronic psychological stress, which negatively affects immune function and overall health status [47]. In summary, the poverty-income ratio plays a partial mediating role in the relationship between depression and mortality through multiple pathways, including economic, psychological, behavioral, and sociostructural factors.

## 5. Limitations

The present study is subject to several limitations that should be acknowledged. Firstly, it should be noted that the data were derived from the National Health and Nutrition Examination Survey (NHANES) conducted in the United States. This may limit the generalizability of the findings to populations in other countries. Secondly, depressive symptoms were only assessed at the baseline, which precluded the analysis of changes in depressive symptoms over time. Furthermore, individuals diagnosed with major depressive disorder may be less inclined to complete the survey or undergo physical examinations due to the severity of their condition. This may result in an underestimation of the true prevalence of depression.

## 6. Conclusion

Based on data from the NHANES, this study found that the famil PIR partially mediates the association between depressive symptoms and the risk of all-cause and cardiovascular mortality. The results indicate that depressive symptoms not only directly increase mortality risk but also further exacerbate adverse outcomes by impacting economic status. Therefore, effective interventions for individuals with depression should address both mental health and socioeconomic support to reduce related mortality risks.

## Data Availability

The datasets used in this study are publicly available from the National Health and Nutrition Examination Survey (NHANES) database (https://www.cdc.gov/nchs/nhanes/index.htm) and the National Death Index (https://www.cdc.gov/nchs/data-linkage/mortality-public.htm).

https://www.cdc.gov/nchs/nhanes/index.htm)?https://www.cdc.gov/nchs/data-linkage/mortality-public.htm

## Notes

### Competing Interest Statement

The authors have declared that no competing interests exist.

### Funding Statement

The author(s) received no specific funding for this work.

### Author Declarations

All participants provided written informed consent, and the study protocol was approved by the NCHS Ethics Review Board. This was a nationally representative cohort study based on NHANES participants.

## References

1. Liu J, Liu Y, Ma W, Tong Y, Zheng J. Temporal and spatial trend analysis of all-cause depression burden based on Global Burden of Disease (GBD) 2019 study. Sci Rep. 2024 May 29;14(1):12346. doi: 10.1038/s41598-024-62381-9. PMID: 38811645; PMCID: PMC11137143.

2. Dettmann LM, Adams S, Taylor G. Investigating the prevalence of anxiety and depression during the first COVID-19 lockdown in the United Kingdom: Systematic review and meta-analyses. Br J Clin Psychol. 2022 Sep;61(3):757–780. doi: 10.1111/bjc.12360. Epub 2022 Feb 9. PMID: 35137427; PMCID: PMC9111383.

3. Chesney E, Goodwin GM, Fazel S. Risks of all-cause and suicide mortality in mental disorders: a meta-review. World Psychiatry. 2014 Jun;13(2):153–60. doi: 10.1002/wps.20128. PMID: 24890068; PMCID: PMC4102288.

4. Lavretsky H, Zheng L, Weiner MW, Mungas D, Reed B, Kramer JH, et al. Association of depressed mood and mortality in older adults with and without cognitive impairment in a prospective naturalistic study. Am J Psychiatry. 2010 May;167(5):589–97. doi: 10.1176/appi.ajp.2009.09020280. Epub 2010 Feb 16. PMID: 20160005; PMCID: PMC2864365.

5. Zhang Z, Jackson SL, Gillespie C, Merritt R, Yang Q. Depressive Symptoms and Mortality Among US Adults. JAMA Netw Open. 2023 Oct 2;6(10):e2337011. doi: 10.1001/jamanetworkopen.2023.37011. PMID: 37812418; PMCID: PMC10562940.

6. Park SH, Kim D, Cho J, Jin Y, Lee I, Lee K, et al. Depressive symptoms and all-cause mortality in Korean older adults: A 3-year population-based prospective study. Geriatr Gerontol Int. 2018 Jun;18(6):950–956. doi: 10.1111/ggi.13270. Epub 2018 Feb 2. PMID: 29392830.

7. Liu T, Wang L, Zhu Z, Wang B, Lu Z, Pan Y, et al. Association of both depressive symptoms scores and specific depressive symptoms with all-cause and cardiovascular disease mortality. Ann Gen Psychiatry. 2024 Jul 15;23(1):25. doi: 10.1186/s12991-024-00509-x. PMID: 39010080; PMCID: PMC11250981.

8. Nabi H, Shipley MJ, Vahtera J, Hall M, Korkeila J, Marmot MG, et al. Effects of depressive symptoms and coronary heart disease and their interactive associations on mortality in middle-aged adults: the Whitehall II cohort study. Heart. 2010 Oct;96(20):1645–50. doi: 10.1136/hrt.2010.198507. Epub 2010 Sep 15. PMID: 20844294; PMCID: PMC3151258.

9. Machado MO, Veronese N, Sanches M, Stubbs B, Koyanagi A, Thompson T, et al. The association of depression and all-cause and cause-specific mortality: an umbrella review of systematic reviews and meta-analyses. BMC Med. 2018 Jul 20;16(1):112. doi: 10.1186/s12916-018-1101-z. PMID: 30025524; PMCID: PMC6053830.

10. Feng Z, Tong WK, Zhang X, Tang Z. Prevalence of depression and association with all-cause and cardiovascular mortality among individuals with type 2 diabetes: a cohort study based on NHANES 2005-2018 data. BMC Psychiatry. 2023 Jul 10;23(1):490. doi: 10.1186/s12888-023-04999-z. PMID: 37430235; PMCID: PMC10331954.

11. Park S, Cho J, Kim D, Jin Y, Lee I, Hong H, et al. Handgrip strength, depression, and all-cause mortality in Korean older adults. BMC Geriatr. 2019 May 3;19(1):127. doi: 10.1186/s12877-019-1140-0. PMID: 31053117; PMCID: PMC6499996.

12. Gathright EC, Goldstein CM, Josephson RA, Hughes JW. Depression increases the risk of mortality in patients with heart failure: A meta-analysis. J Psychosom Res. 2017 Mar;94:82–89. doi: 10.1016/j.jpsychores.2017.01.010. Epub 2017 Jan 24. PMID: 28183407; PMCID: PMC5370194.

13. Lee J, Choi KS, Yun JA. The effects of sociodemographic factors on help-seeking for depression: Based on the 2017-2020 Korean Community Health Survey. PLoS One. 2023 Jan 19;18(1):e0280642. doi: 10.1371/journal.pone.0280642. PMID: 36656907; PMCID: PMC9851551.

14. Mehta KM, Yaffe K, Langa KM, Sands L, Whooley MA, Covinsky KE. Additive effects of cognitive function and depressive symptoms on mortality in elderly community-living adults. J Gerontol A Biol Sci Med Sci. 2003 May;58(5):M461-7. doi: 10.1093/gerona/58.5.m461. PMID: 12730257; PMCID: PMC2939722.

15. Eriksson MD, Eriksson JG, Korhonen P, Koponen H, Salonen MK, Mikkola TM, et al. Depressive symptoms and mortality-findings from Helsinki birth cohort study. Acta Psychiatr Scand. 2023 Feb;147(2):175–185. doi: 10.1111/acps.13512. Epub 2022 Nov 1. PMID: 36263580; PMCID: PMC10092352.

16. Li F, Chu Z. The protective role of employment in depression: insights from 2005 to 2018 NHANES information. Front Psychiatry. 2024 Dec 9;15:1455122. doi: 10.3389/fpsyt.2024.1455122. PMID: 39720425; PMCID: PMC11667109.

17. Lorant V, Deliège D, Eaton W, Robert A, Philippot P, Ansseau M. Socioeconomic inequalities in depression: a meta-analysis. Am J Epidemiol. 2003 Jan 15;157(2):98–112. doi: 10.1093/aje/kwf182. PMID: 12522017.

18. Lund C, Breen A, Flisher AJ, Kakuma R, Corrigall J, Joska JA, et al. Poverty and common mental disorders in low and middle income countries: A systematic review. Soc Sci Med. 2010 Aug;71(3):517–528. doi: 10.1016/j.socscimed.2010.04.027. Epub 2010 May 12. PMID: 20621748; PMCID: PMC4991761.

19. Flèche S, Layard R. Do More of Those in Misery Suffer from Poverty, Unemployment or Mental Illness? Kyklos (Oxford). 2017 Feb;70(1):27–41. doi: 10.1111/kykl.12129. Epub 2017 Jan 21. PMID: 28729747; PMCID: PMC5511887.

20. Li Y, Chen T, Li Q, Jiang L. The Impact of Subjective Poverty on the Mental Health of the Elderly in China: The Mediating Role of Social Capital. Int J Environ Res Public Health. 2023 Aug 29;20(17):6672. doi: 10.3390/ijerph20176672. PMID: 37681812; PMCID: PMC10487636.

21. Ssebunnya J, Kigozi F, Lund C, Kizza D, Okello E. Stakeholder perceptions of mental health stigma and poverty in Uganda. BMC Int Health Hum Rights. 2009 Mar 31;9:5. doi: 10.1186/1472-698X-9-5. PMID: 19335889; PMCID: PMC2670268.

22. Kroenke K, Spitzer RL, Williams JB. The PHQ-9: validity of a brief depression severity measure. J Gen Intern Med. 2001 Sep;16(9):606–13. doi: 10.1046/j.1525-1497.2001.016009606.x. PMID: 11556941; PMCID: PMC1495268.

23. Negeri ZF, Levis B, Sun Y, He C, Krishnan A, Wu Y, et al. Thombs BD; Depression Screening Data (DEPRESSD) PHQ Group. Accuracy of the Patient Health Questionnaire-9 for screening to detect major depression: updated systematic review and individual participant data meta-analysis. BMJ. 2021 Oct 5;375:n2183. doi: 10.1136/bmj.n2183. PMID: 34610915; PMCID: PMC8491108.

24. Manea L, Gilbody S, McMillan D. A diagnostic meta-analysis of the Patient Health Questionnaire-9 (PHQ-9) algorithm scoring method as a screen for depression. Gen Hosp Psychiatry. 2015 Jan-Feb;37(1):67-75. doi: 10.1016/j.genhosppsych.2014.09.009. Epub 2014 Sep 23. PMID: 25439733..

25. Johnson CJ, Weir HK, Fink AK, German RR, Finch JL, Rycroft RK, et al. Accuracy of Cancer Mortality Study Group. The impact of National Death Index linkages on population-based cancer survival rates in the United States. Cancer Epidemiol. 2013 Feb;37(1):20–8. doi: 10.1016/j.canep.2012.08.007. Epub 2012 Sep 7. PMID: 22959341; PMCID: PMC4558883.

26. Huang Y, Chen X, Cai X. The non-linear association between depression scores and all-cause mortality: a cohort study based on NHANES 2005-2018 data. Sci Rep. 2025 May 3;15(1):15492. doi: 10.1038/s41598-025-00366-y. PMID: 40319084; PMCID: PMC12049539.

27. Hansel B, Potier L, Chalopin S, Larger E, Gautier JF, Delestre F, et al. The COVID-19 lockdown as an opportunity to change lifestyle and body weight in people with overweight/obesity and diabetes: Results from the national French COVIDIAB cohort. Nutr Metab Cardiovasc Dis. 2021 Aug 26;31(9):2605–2611. doi: 10.1016/j.numecd.2021.05.031. Epub 2021 Jun 17. PMID: 34348875; PMCID: PMC9187903.

28. World Health Organization (2000) Obesity: Preventing and Managing the Global Epidemic. Report of a WHO Consultation. WHO Technical Report Series 894. Geneva: World Health Organization. Available: https://www.who.int/nutrition/publications/obesity/WHO_TRS_894/en/

29. Imai K, Keele L, Tingley D. A general approach to causal mediation analysis. Psychol Methods. 2010 Dec;15(4):309–34. doi: 10.1037/a0020761. PMID: 20954780.

30. Wang Z, Pu B. Joint effects of depression and social determinants of health on mortality risk among U.S. adults: a cohort study. BMC Psychiatry. 2024 Oct 30;24(1):752. doi: 10.1186/s12888-024-06159-3. PMID: 39478508; PMCID: PMC11523881.

31. Pratt LA, Druss BG, Manderscheid RW, Walker ER. Excess mortality due to depression and anxiety in the United States: results from a nationally representative survey. Gen Hosp Psychiatry. 2016 Mar-Apr;39:39-45. doi: 10.1016/j.genhosppsych.2015.12.003. Epub 2015 Dec 18. PMID: 26791259; PMCID: PMC5113020.

32. Brandão DJ, Fontenelle LF, da Silva SA, Menezes PR, Pastor-Valero M. Depression and excess mortality in the elderly living in low- and middle-income countries: Systematic review and meta-analysis. Int J Geriatr Psychiatry. 2019 Jan;34(1):22–30. doi: 10.1002/gps.5008. Epub 2018 Nov 15. PMID: 30306638.

33. Falagas ME, Vouloumanou EK, Mavros MN, Karageorgopoulos DE. Economic crises and mortality: a review of the literature. Int J Clin Pract. 2009 Aug;63(8):1128–35. doi: 10.1111/j.1742-1241.2009.02124.x. PMID: 19624782.

34. Sabanayagam C, Shankar A. Income is a stronger predictor of mortality than education in a national sample of US adults. J Health Popul Nutr. 2012 Mar;30(1):82–6. doi: 10.3329/jhpn.v30i1.11280. PMID: 22524123; PMCID: PMC3312363.

35. Hone T, Mirelman AJ, Rasella D, Paes-Sousa R, Barreto ML, Rocha R, et al. Effect of economic recession and impact of health and social protection expenditures on adult mortality: a longitudinal analysis of 5565 Brazilian municipalities. Lancet Glob Health. 2019 Nov;7(11):e1575–e1583. doi: 10.1016/S2214-109X(19)30409-7. PMID: 31607469.

36. Han RH, Schmidt MN, Waits WM, Bell AKC, Miller TL. Planning for Mental Health Needs During COVID-19. Curr Psychiatry Rep. 2020 Oct 8;22(12):66. doi: 10.1007/s11920-020-01189-6. PMID: 33030637; PMCID: PMC7542088.

37. Shukla J, Manohar Singh R. Psychological Health amidst COVID-19: A Review of existing literature in the Indian Context. Clin Epidemiol Glob Health. 2021 Jul-Sep;11:100736. doi: 10.1016/j.cegh.2021.100736. Epub 2021 Apr 17. PMID: 33898865; PMCID: PMC8052584.

38. Garcini LM, Rosenfeld J, Kneese G, Bondurant RG, Kanzler KE. Dealing with distress from the COVID-19 pandemic: Mental health stressors and coping strategies in vulnerable latinx communities. Health Soc Care Community. 2022 Jan;30(1):284–294. doi: 10.1111/hsc.13402. Epub 2021 Apr 24. PMID: 33894080; PMCID: PMC8251305.

39. Huttunen-Lenz M, Raben A, Adam T, Macdonald I, Taylor MA, Stratton G, et al. Socio-economic factors, mood, primary care utilization, and quality of life as predictors of intervention cessation and chronic stress in a type 2 diabetes prevention intervention (PREVIEW Study). BMC Public Health. 2023 Aug 30;23(1):1666. doi: 10.1186/s12889-023-16569-9. PMID: 37649005; PMCID: PMC10466828.

40. Starnes JR, Di Gravio C, Irlmeier R, Moore R, Okoth V, Rogers A, et al. Characterizing multidimensional poverty in Migori County, Kenya and its association with depression. PLoS One. 2021 Nov 16;16(11):e0259848. doi: 10.1371/journal.pone.0259848. PMID: 34784390; PMCID: PMC8594838.

41. Wickrama KA, Surjadi FF, Lorenz FO, Conger RD, Walker C. Family Economic Hardship and Progression of Poor Mental Health in Middle-aged Husbands and Wives. Fam Relat. 2012 Apr 1;61(2):297–312. doi: 10.1111/j.1741-3729.2011.00697.x. Epub 2012 Mar 13. PMID: 22577243; PMCID: PMC3346274.

42. Pulgar CA, Trejo G, Suerken C, Ip EH, Arcury TA, Quandt SA. Economic Hardship and Depression Among Women in Latino Farmworker Families. J Immigr Minor Health. 2016 Jun;18(3):497–504. doi: 10.1007/s10903-015-0229-6. PMID: 26022147; PMCID: PMC4663186.

43. Ridley M, Rao G, Schilbach F, Patel V. Poverty, depression, and anxiety: Causal evidence and mechanisms. Science. 2020 Dec 11;370(6522):eaay0214. doi: 10.1126/science.aay0214. Erratum in: Science. 2024 Mar 22;383(6689):eadp1916. doi: 10.1126/science.adp1916. PMID: 33303583.

44. Stevenson C, Wakefield JRH, Bowe M, Kellezi B, Jones B, McNamara N. Weathering the economic storm together: Family identification predicts future well-being during COVID-19 via enhanced financial resilience. J Fam Psychol. 2022 Apr;36(3):337–345. doi: 10.1037/fam0000951. Epub 2022 Jan 24. PMID: 35073125.

45. Menec VH, Newall NE, Mackenzie CS, Shooshtari S, Nowicki S. Examining individual and geographic factors associated with social isolation and loneliness using Canadian Longitudinal Study on Aging (CLSA) data. PLoS One. 2019 Feb 1;14(2):e0211143. doi: 10.1371/journal.pone.0211143. PMID: 30707719; PMCID: PMC6358157.

46. Akgül H, Güven AZ, Güven S, Ceylan M. Loneliness, Social Support, Social Trust, and Subjective Wellness in Low-Income Children: A Longitudinal Approach. Children (Basel). 2023 Aug 23;10(9):1433. doi: 10.3390/children10091433. PMID: 37761396; PMCID: PMC10529055.

47. Rutakumwa R, Tusiime C, Mpango RS, Kyohangirwe L, Kaleebu P, Patel V, et al. A Qualitative Exploration of Causes of Depression among Persons Living with HIV Receiving Antiretroviral Therapy in Uganda: Implications for Policy. Psychiatry J. 2023 Jan 17;2023:1986908. doi: 10.1155/2023/1986908. PMID: 36704236; PMCID: PMC9873437.

48. Lago-Hernandez C, Nguyen NH, Khera R, Loomba R, Asrani SK, Singh S. Financial Hardship From Medical Bills Among Adults With Chronic Liver Diseases: National Estimates From the United States. Hepatology. 2021 Sep;74(3):1509–1522. doi: 10.1002/hep.31835. Epub 2021 Jun 2. PMID: 33772833.

49. Jebena MG, Lindstrom D, Belachew T, Hadley C, Lachat C, Verstraeten R, et al. Food Insecurity and Common Mental Disorders among Ethiopian Youth: Structural Equation Modeling. PLoS One. 2016 Nov 15;11(11):e0165931. doi: 10.1371/journal.pone.0165931. PMID: 27846283; PMCID: PMC5113011.

50. Saito M, Kondo K, Kondo N, Abe A, Ojima T, Suzuki K; JAGES group. Relative deprivation, poverty, and subjective health: JAGES cross-sectional study. PLoS One. 2014 Oct 28;9(10):e111169. doi: 10.1371/journal.pone.0111169. PMID: 25350284; PMCID: PMC4211701.

51. Hatcher AM, Gibbs A, Jewkes R, McBride RS, Peacock D, Christofides N. Effect of Childhood Poverty and Trauma on Adult Depressive Symptoms Among Young Men in Peri-Urban South African Settlements. J Adolesc Health. 2019 Jan;64(1):79–85. doi: 10.1016/j.jadohealth.2018.07.026. Epub 2018 Oct 14. PMID: 30327276.

